# Global and Historical Distribution of *Clostridioides difficile* in the Human Diet (1981-2019): Systematic Review and Meta-Analysis of 21886 Samples Reveal Sources of Heterogeneity, High-Risk Foods, and Unexpected Higher Prevalence Towards the Tropic

**DOI:** 10.1101/19012450

**Authors:** Alexander Rodriguez-Palacios, Kevin Q Mo, Bhavan U. Shah, Joan Msuya, Nina Bijedic, Abhishek Deshpande, Sanja Ilic

**Affiliations:** Division of Gastroenterology and Liver Disease, Case Western Reserve University School of Medicine, Cleveland, OH 44106, USA; Digestive Health Research Institute, Case Western Reserve University School of Medicine, Cleveland, OH 44106, USA; Department of Human Sciences, Human Nutrition, College of Education and Human Ecology, The Ohio State University, Columbus, OH 43210, USA; Northeast Ohio University of Medicine, Rootstown, OH 44272, USA; Informatics and Assessment Division, Lorain County General Health District, Elyria, OH 44035, USA; Department of Neurology, Weill Cornell Medicine, Cornell University, New York, NY 10065, USA; Department of Health and Nutrition, World Vision, Arusha, Tanzania; Department of Applied Mathematics and Formal Methods, Information Technologies, University Dzemal Bijedic, Mostar 88000, Bosnia and Herzegovina; Department of Mathematics, University of North Carolina, Charlotte, NC 28262, USA; Medicine Institute Center for Value-Based Care Research, Cleveland Clinic, Cleveland, OH 44106 USA

**Author notes:** **Correspondence:** (AR-P), (SI). **Authors’ Contribution** SI and ARP designed the study. SI and AD developed the search algorithm; SI, JM, ARP, and BS developed and pretested data extraction strategy and tool. JM BS, KM, ARP, and JM performed the literature search. KM and BS under the supervision of SI and AR extracted the data. AR SI planned and performed all data analysis with support from NB. SI and ARP wrote the manuscript. All authors read and approved submission of this manuscript.

## Abstract

*Clostridioides difficile* (CD) is a spore-forming bacterium that causes life-threatening intestinal infections in humans. Although formerly regarded as exclusively nosocomial, there is increasing genomic evidence that person-to-person transmission accounts for only <25% of cases, supporting the culture-based hypothesis that foods may be routine sources of CD-spore ingestion in humans.

To synthesize the evidence on the risk of CD exposure via foods, we conducted a systematic review and meta-analysis of studies reporting the culture prevalence of CD in foods between January 1981 and November 2019. Meta-analyses, risk-ratio estimates, and meta-regression were used to estimate weighed-prevalence across studies and food types to identify laboratory and geographical sources of heterogeneity.

In total, 21,886 food samples were tested for CD between 1981 and 2019 (232 food-sample-sets; 79 studies; 25 countries). Culture methodology, sample size and type, region, and latitude were significant sources of heterogeneity (p<0.05). Although non-strictly-anaerobic methods were reported in some studies, and we confirmed experimentally that improper anaerobiosis of media/sample-handling affects CD recovery in agar (Fisher, p<0.01), most studies (>72%) employed the same (one-of-six) culture strategy. Because the prevalence was also meta-analytically similar across six culture strategies reported, all studies were integrated using three meta-analytical methods. At the study level (n=79), the four-decade global cumulative-prevalence of CD in the human diet was 4.1% (95%CI=-3.71, 11.91). At the food-set level (n=232), the weighted prevalence ranged between 4.5% (95%CI=3-6%; all studies) and 8% (95%CI=7-8%; only CD-positive-studies). Risk-ratio ranking and meta-regression showed that milk was the least likely source of CD, while seafood, leafy green vegetables, pork, and poultry carried higher risks (p<0.05). Across regions, the risk of CD in foods for foodborne exposure reproducibly decreased with Earth latitude (p<0.001).

In conclusion, CD in the human diet is a global nonrandom-source of foodborne exposure that occurs independently of laboratory culture methods, across regions, and at variable level depending on food type and latitude. The latitudinal trend (high CD-food-prevalence towards tropic) is unexpectedly inverse to the epidemiological observations of CD-infections in humans (frequent in temperate regions). Findings suggests the plausible hypothesis that ecologically-richer microbiomes in the tropic might protect against intestinal CD colonization/infections despite CD ingestion.

## Introduction

*Clostridioides* (*Clostridium) difficile* (CD) is a spore-forming anaerobic bacterium that causes severe enteritis, colitis, and mortality in susceptible humans, especially if affected with inflammatory bowel diseases, cancer, immunosuppression, or if taking antibiotics.^110-1213^ To date, it is well known that CD infections (CDI) in humans are more frequent in temperate regions. Latitudinal trends however have not been reported for CDI at continental scales. Since the first report linking CD to pseudomembranous colitis in 1975, several reports now indicate that CD could reach humans via foods.^2^ If the presence of *C. difficile* in foods was indeed linearly associated with infections, one would expect that the prevalence of food contamination was higher in temperate regions as it is the case for the incidence of CDI in humans.

CDI have now worsened severity and incidence since the emergence of hypervirulent strains that caused CDI epidemics in both Canada and the UK in the mid 2000s. After the astounding isolation of such strains from young cattle and retail beef in Canada in 2005 ^3,4^ numerous food studies support the hypothesis of potential foodborne exposure ^5,6^. With the availability of genomics, elegant studies have shown that only ∼25-30% of CDI in hospitals are nosocomial, redirecting the attention to foods as viable sources of CD. ^89^ As further evidence for connectivity between foods and CDI, last year a *de novo* genome sequencing study showed that the first CD strain derived from foods (PCR ribotype 078) in Canada in 2005 was identical to the historical strain M120 that contributed to epidemics in the UK in 2007 ^7^.

Unless we understand the distribution pattern of CD across foods, regions, and laboratory variability, little can be done to minimize the exposure of susceptible persons to CD in their diet. Distinguishing methodological variability from natural variability is important to assign a proper risk value to the presence of CD in the food supply ^2^. To formally quantify the prevalence of CD in foods and map the distributional trends over global scales, we conducted a systematic review and meta-regression of studies reporting the presence of CD in foods. The main quantitative objectives were *i)* to appraise peer-reviewed studies on quality and the prevalence of CD in foods, *ii)* to determine laboratory factors associated with CD-positivity, and iii) to perform meta-analysis across regions, and food items to examine reporting differences and outline latitudinal trends.

Herein, we report that the majority of studies used the same laboratory culture method for the isolation of CD allowing us to conduct meta-analysis and rank food items based on the weighted risk of contamination across regions. Although beef and pork were food categories often containing CD, leafy green vegetables and seafoods had higher rates of contamination. Of remarkable novelty, the contamination of foods followed a latitudinal trend that is inverse to the Earth’s latitude (higher towards the Equator). Although there are no global reports describing latitudinal trends for CDI, results indicate that the latitudinal trend observed in foods is inverse to that of what is reported and expected for infections (*i.e*., high incidence in temperate regions).

### Materials and Methods

#### Systematic Review, Team and Definitions

This study follows and complies with principles of systematic review research methodology for ‘food safety’ and food item definitions.^14,15^ All procedures used in this study were reported in accordance with Preferred Reporting Items for Systematic Reviews and Meta-Analyses (PRISMA) guidelines in structuring our literature search analysis.

We conducted a systematic search of available literature reporting the prevalence of *C. difficile* in foods. Electronic databases (MEDLINE/PubMed, Scopus, Web of Science, and Google Scholar) were searched to identify all studies reporting the prevalence of CD in foods. The detailed search algorithm, questionnaire, data extraction criteria and verification are available as **Supplementary Materials**. Five iterative rounds of verification of extraction strategies and tools were validated to ensure reproducibility of data extraction.

In brief, a list of search terms was developed by consensus by the research team to retrieve citations pertaining to CD prevalence in foods. Search terms (n= 64 terms) relating to population (*e.g*. food, meat, beef, etc.) and outcome (*e.g*., *Clostridium Clostridioides difficile*) were combined to search numerous food types (or items), without restrictions. Identified terms were pre-tested in PubMed and used to develop the final algorithm, using as basis a similar validated strategy we implemented for vegetables. ^19^ The complete search terms consisted of the following: “(*C. difficile* OR *Clostridium difficile* OR *difficile*) AND (food* OR meal* OR mollus* OR fish OR crustaceans OR oysters OR poultry OR chicken OR turkey OR duck OR goose OR meat* OR beef OR pork OR venison OR dairy OR milk OR yogurt OR cheese OR egg* OR sausage OR seafood OR butter OR lard OR honey OR vegetable* OR lettuce OR spinach OR cabbage OR fresh leafy green herb* OR endive OR arugula OR chard OR watercress OR radicchio OR frieze OR mustard green OR beans OR cauliflower OR broccoli OR celery OR onion OR cantaloupe OR watermelon OR melon OR mushroom OR carrot OR potato OR garlic OR radish OR corn OR peas OR cucumber OR tomato OR pepper* OR alfalfa OR sprout*)”, and was used to search Web of Science, Scopus Cochrane and Pub Med. The search was repeated regularly and database updated until the last update in November 2019, prior to the manuscript submission.

Citations retrieved from electronic databases were imported and de-duplicated in reference management software EndNote Web™ (Clarivate Analytics). Search verification included manual searching of references citing the first five manuscripts reporting CD in foods or its potential for foodborne transmission ^9, 10, 26, 44, 45^ using Google Scholar in consultation with research team members, and the references of all identified studies. Experts in the field were consulted to identify unpublished data, including theses and research poster/conference presentations. Google Search Engine limited to the first 600 hits was searched to identify any “gray literate”. Alert in Google Scholar was set up to identify any newly published studies. All potentially relevant citations discovered through the manual searching method, which were not previously identified through electronic search, were added into the review process and processed in the same manner as electronic citations. All peer-reviewed studies, dissertations and reports containing original prevalence data were eligible. Studies lacking the report of both number of samples tested (*N*) and number of positive samples (*n*) were excluded. ^32^ Prevalence contamination data was only extracted for culture assays, and not for prevalence data based on molecular assays. ^27^ No restrictions were imposed in terms of the study time period, design, language, or study origin.

Relevant citations after reviewer screen 1 (RS1) were procured as full articles, and screened by two reviewers (BS and SI) using pre-tested RS2 checklists (**Supplementary Tables 1** and **2**). Conflicts were resolved by a consensus between respective reviewers and when not possible, by senior authors of this study. During initial manual screening of selected abstracts, carcass trims or carcass washings/rinsates at the processing plants were selected for secondary analysis. Data describing environment, wastewater, animal or human fecal samples were excluded. Non-primary research studies (e.g. narrative reviews) and studies investigating other aspects (e.g. outbreak reports, test performance studies) were excluded. Case reports or case series of hospital-associated *C. difficile* infections, and case-control studies that did not provide prevalence estimates, and duplicate publications were also excluded. Relevant articles were assessed and categorized by food type (e.g. beef, poultry, vegetables) and descriptive characteristics (e.g., food processing level, where in the production chain was the product sampled). Through initial title and abstract-based relevance screening one (RS1), potentially relevant primary research articles were identified.

#### Extraction Tool and Risk of Bias

Prior to reading the manuscripts, two meeting sessions (phone, and in person) took place (ARP, SI, BS, and AD) to discuss and create a Data Extraction Tool (DET, list of questions and response categories, see **Supplementary Table 1**) draft to standardize the extraction of data required for statistical analysis and testing of study objectives. Following five iterative rounds of verification for accuracy and clarity, the pre-final extraction tool was pretested by ARP and BS at CWRU, and SI and JM at OSU, using 10 studies (the first five in the 1980s, and 5 in 2015) ^9, 10, 26, 44, 45^. Phone conferences occurred biweekly during this phase to estimate test agreement, to address concerns, to edit/improve, and thus finalize the DET. The pretesting and definitive data extraction were conducted after the participating reviewers (KM, BS) were trained on laboratory methodologies available for CD by senior scientists from two institutions (ARP and SI). Four reviewers extracted data independently. Data extraction was verified by senior two authors for data interpretation, extraction and accuracy. The final DET was used to extract all relevant research articles, which were assessed for methodological soundness and bias as part of the data extraction strategy, by at least two reviewers, using the prevalence study-based criteria.

All studies were assessed by rating each of the 6 quality assessment items listed in the DET into dichotomous ratings: low risk (1) and high risk (0). An overall Risk of Bias score was calculated by adding the numeric value of all six items. High scores indicate low risk of bias and stronger method quality.

Measures of data SD or variability were estimated using the number of food samples tested and the percentage of positive samples. Because the reliability of available statistical methods on bias have previously shown to be inaccurate and misleading with effects that are close to the extremes, for instance close to 0 or to 100% ^24^, publication bias was tested using funnel plot and Egger’s statistics using study size at the food set level instead of the standard error of the effect as recommended for proportions with high data /effects polarity ^24^. As we recently mentioned ^25^ however, it is uncertain how many studies start but do not get published due to the lack of a prepublication registry of prevalence based studies in foods.

#### Pooled Ratios, Meta-analysis and Meta-regression

Extracted data were used to estimate risk ratios and perform a prevalence meta-analysis. Three main categories of data were extracted: sample characteristics, methods, and prevalence data. All food items were grouped for analytical purposes into food item categories (*e.g*., pork, leafy green vegetables). Pooled risk ratios (RR, 95% CI) for each food group were calculated to quantify the differences and rank the foods according to the risk of being contaminated using a random effects model ^2618^ In brief, heterogeneity tests with Higgins’ I^2^ statistic were performed to determine the extent of variation between the studies that rely on measure analysis for the deviations for each within-study variance from a central estimate for the collective between-study variance distribution. ^26^ Meta-analysis was used to estimate the overall prevalence of CD in foods globally and per region by pooling variances of proportions in a random-effects model using DerSimonian and Laird method. ^28,29^ Analyses were performed using R software and *Metaphor* ^33^, and Stata’s *Metaregression and Metacum* functions. To illustrate the cumulative meta-analytical prevalence of CD globally and regionally at the ‘study-level’ (n=79), over the past 4 decades, we analyzed and plotted the data as a forest plot as previously reported ^25^.

Because each study tested multiple ‘food item categories’, we then decomposed the study variance across each food item, within each study, and constructed the remaining forest plots at the item level presented in this study. Exact binomial weighted and pooled estimates at ‘item level’ (n=232) are presented in forest plots both without adjusting for ‘zero-studies’ (which excludes 0% prevalence studies), and with adjustments using either a balanced addition of 1 to n and N, or using the Freeman-Tukey double arcsine transformation, which include 0% prevalence studies. ^33^ For meta-regression and latitudinal analysis, coordinate data were obtained from NASA. To determine if the reported prevalence was influenced by the amount of food tested, data were extracted as absolute values in grams. Modeling and latitudinal simulations were conducted in R and STATA ^33^ (**Supplementary Materials**).

#### Experiments with *C. difficile* on non-anaerobic media

The exposure of CD spores to conditions suitable for grow (high moisture, nutrients, and warmth) trigger spore germination even in room air. However, the subsequent step, *i.e*., bacterial growth from germinated spore to cell division does not occur in the presence of air/oxygen. Because ***i)*** most studies did not report whether the reagents or the handling of foods in growth media were fully anaerobically, and because ***ii)*** the germination of CD spores and the subsequent viability of vegetative daughter cells are influenced by the lack of strict anaerobiosis, we determined if a source of low CD recovery and study variability could be partly due to negative selection when non-reduced reagents are used. To test this hypothesis we platted 1-year old (superdormant) spores aged for 1 year in PBS as described ^36^ on TSA agar enriched with 5% defibrinated sheep blood. Two different pre-reduced agar conditions, which only differed on the length of time the agar had been incubated (pre-reduced) anaerobically before being used for bacterial inoculation using our Parallel Lanes Plating method.^37^

#### Authorship, Writing and Editorial Assistance

All authors meet the International Committee of Medical Journal Editors (ICMJE) criteria for authorship for this article, take responsibility for the integrity of the work as a whole, and have given their approval for this version to be published. The statistical analysis, interpretation and writing of the manuscript was conducted by the authors without the aid of fee-for-service commercial services.

#### Disclosures and Compliance with Ethics

The authors declare that there is no conflict of interests regarding the publication of this paper. A.D. has received research support from Clorox and is on the advisory board of Ferring Pharmaceuticals, unrelated to the study. This article is based on previously conducted studies, including publications authored by ARP and SI, but does not contain unpublished studies performed by any of the authors.

#### Data Availability and Open Access

All data, easily inferable from the figures and supplementary materials and which is also easily extractable from the manuscripts listed in the **Supplementary References**, will be made fully available by the corresponding authors upon reasonable request. This article is intended for publication as open access under the terms of a Creative Commons Attribution-International License.

## Results

### Global Distribution of Studies Reporting *C. difficile* in Foods and Publication Bias

The PRISMA-standard **Figure 1** summarizes the process of studies selection for this systematic review. From 1,939 studies identified, 79 fulfilled the inclusion criteria for meta-analysis, involving >231 authors and 25 countries over the past 37 years. The list of studies in chronological order from the Americas ^6,18,19,34,35,38-67^, Europe ^23,68-88^, Asia ^89-103^, Africa ^104-109^, and Oceania (Australia/New Zealand) ^20,110,111^ is presented in **Table 1**. Eleven (13.9%) of all studies reported the absence of CD in the food samples tested (CD-negative). Only 30% of studies (n=24) were dedicated to testing only one type of food. Most studies tested between 2 and 4 food types. Funnel plot analysis indicate there has been absent-to-moderate publication bias, depending on the statistical method used for the funnel analysis to consider data handling of reports close to 0% prevalence as illustrated in **Figure 2**.

**Table 1.**
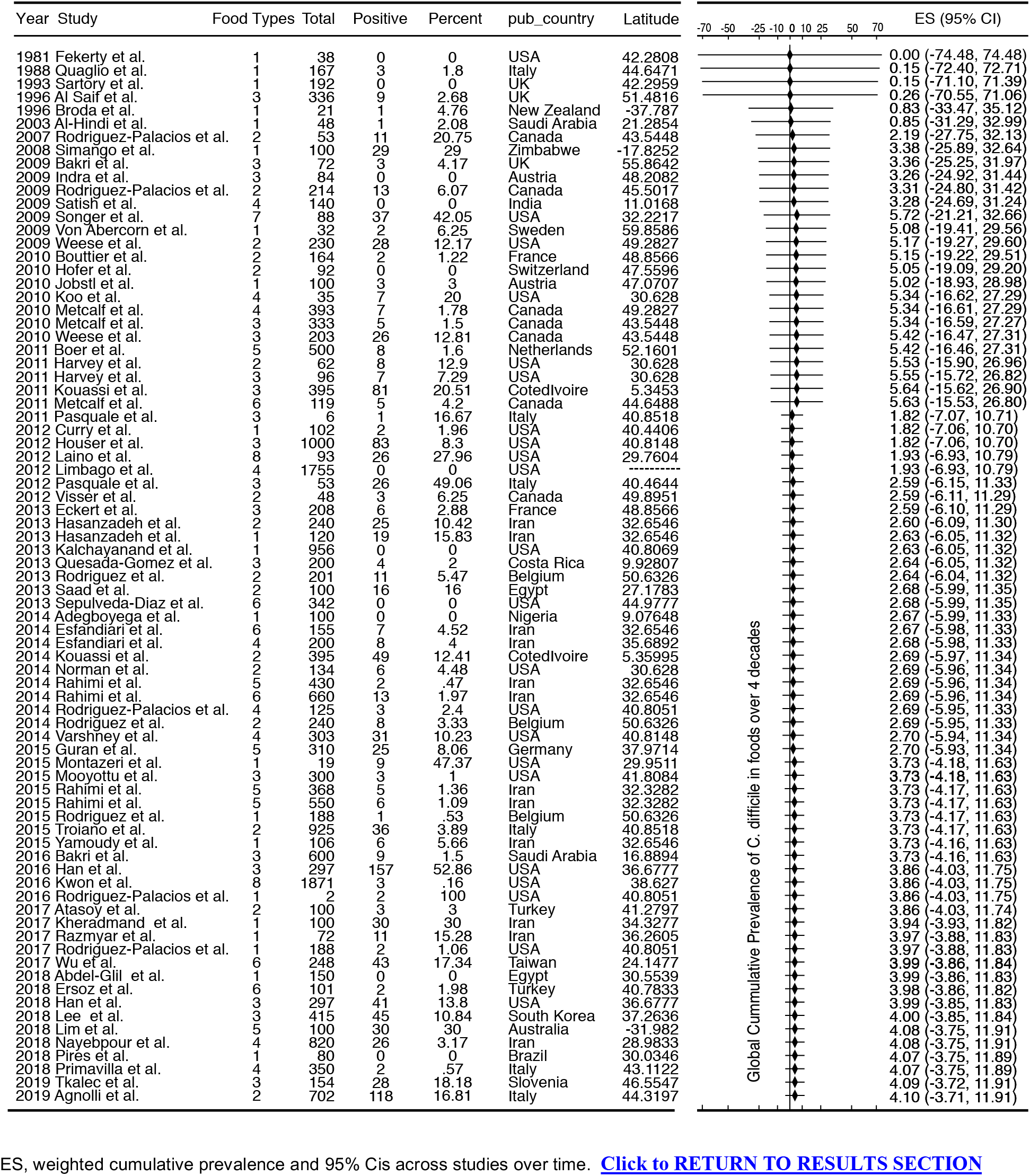
List of studies included in this meta-analysis and their collective cumulative prevalence.

**Figure 1.**
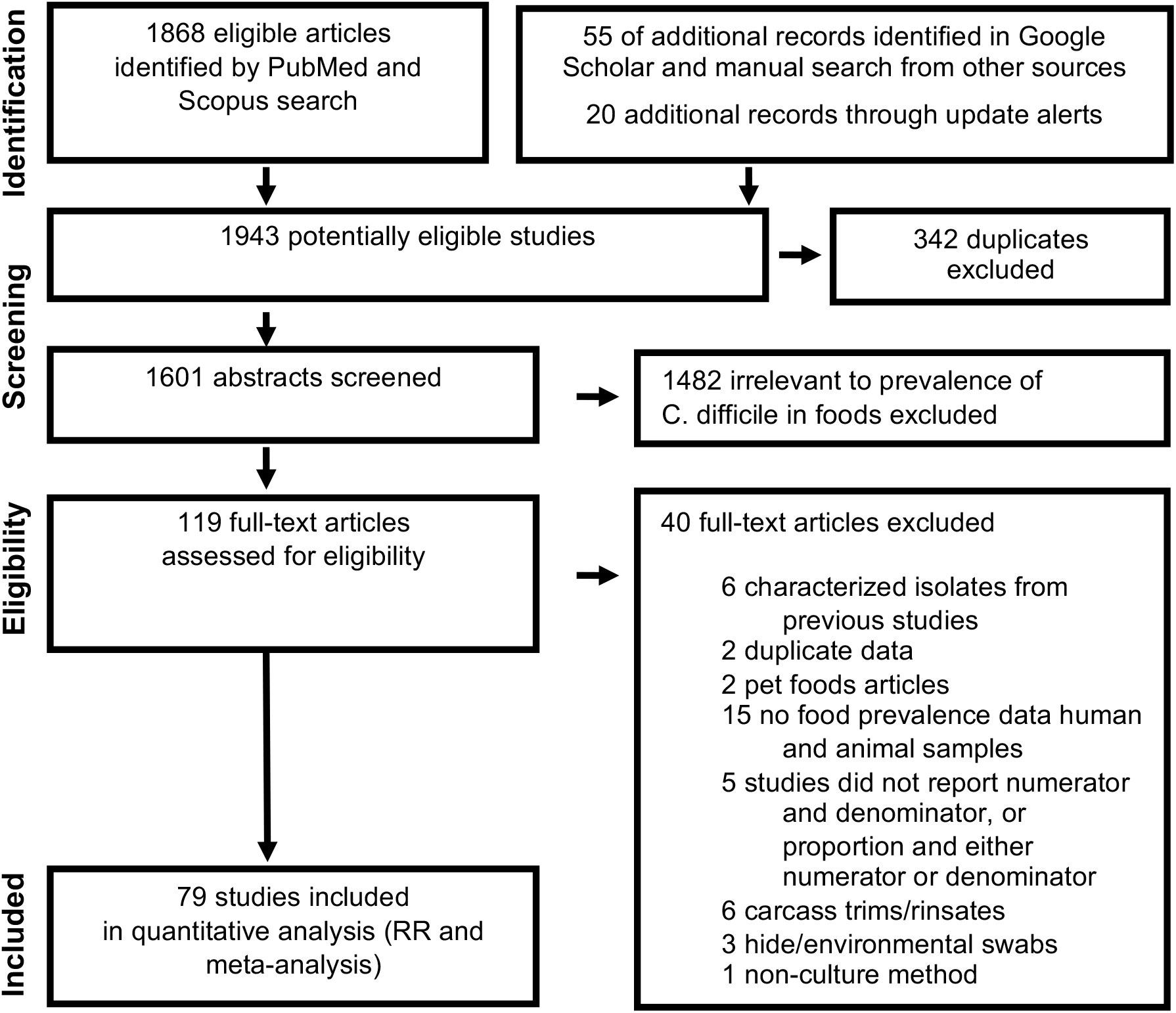
PRISMA selection of studies reporting ‘*C difficile* prevalence in foods’ included in this meta-analysis. The final dataset includes data from 232 food item sample sets reported in 79 studies (Table 1 and Figures 1 and 2 illustrate the distribution of studies conducted in America, ^1-5, 7-30, 79-84^, Europe, ^31-35, 37-53^ Asia, ^54-67, 70^ Africa, ^71-76^ and Oceania,^77, 78, 85^. References for all included studies are available in **Supplementary Materials.**

**Figure 2.**
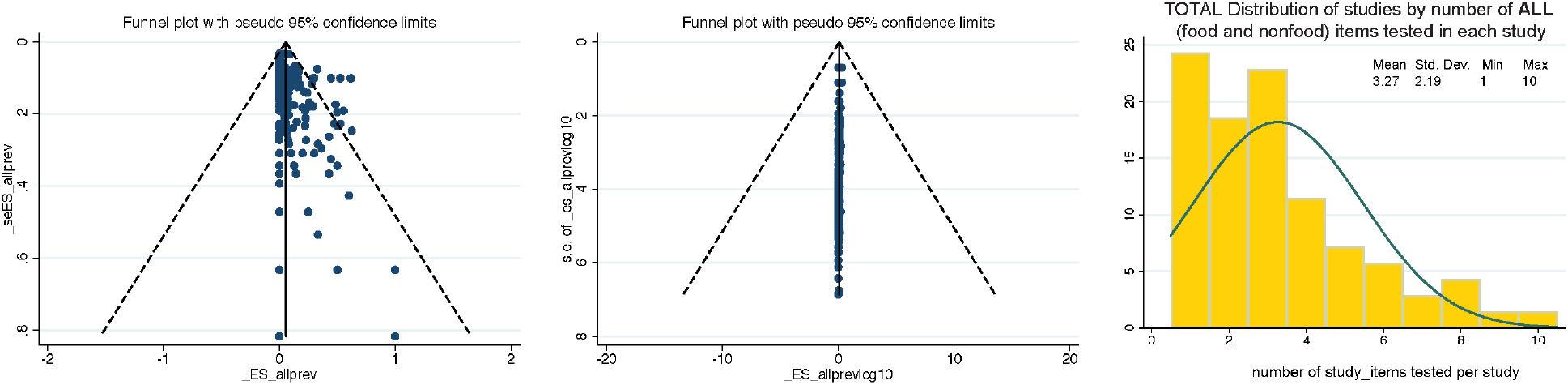
Funnel plot analysis of *C. difficile* prevalence reports and distribution of studies by the number of food items tested per study. **a**) Funnel plots for bias. Panel on the left is the standard plot of weighted estimated prevalence vs. standard error of estimated prevalence, however, such strategy is misleading towards suggesting there is publication bias when used on proportion based meta-analyses if the reported effects are close to zero^86^. The alternative panel to the right is weighted estimated prevalence vs sample size^86^. **b**) Histogram. Study distributions categorized based on total number of food/nonfood item categories tested in each study.

### Historical Study Referents of *C. difficile* Isolation from Foods

This meta-analysis illustrates the geographical distributions of the numerous laboratories around the world that have been examining the potential of foodborne transmissibility of CD spores to humans, via the food supply. **Figure 3** depicts in a map the arithmetic average of the CD prevalence reported for local food items across countries, and other descriptive features of the studies. Of note, since the first report, there have been periods of oscillations possibly reflecting trends in research interest or funding availability.

**Figure 3.**
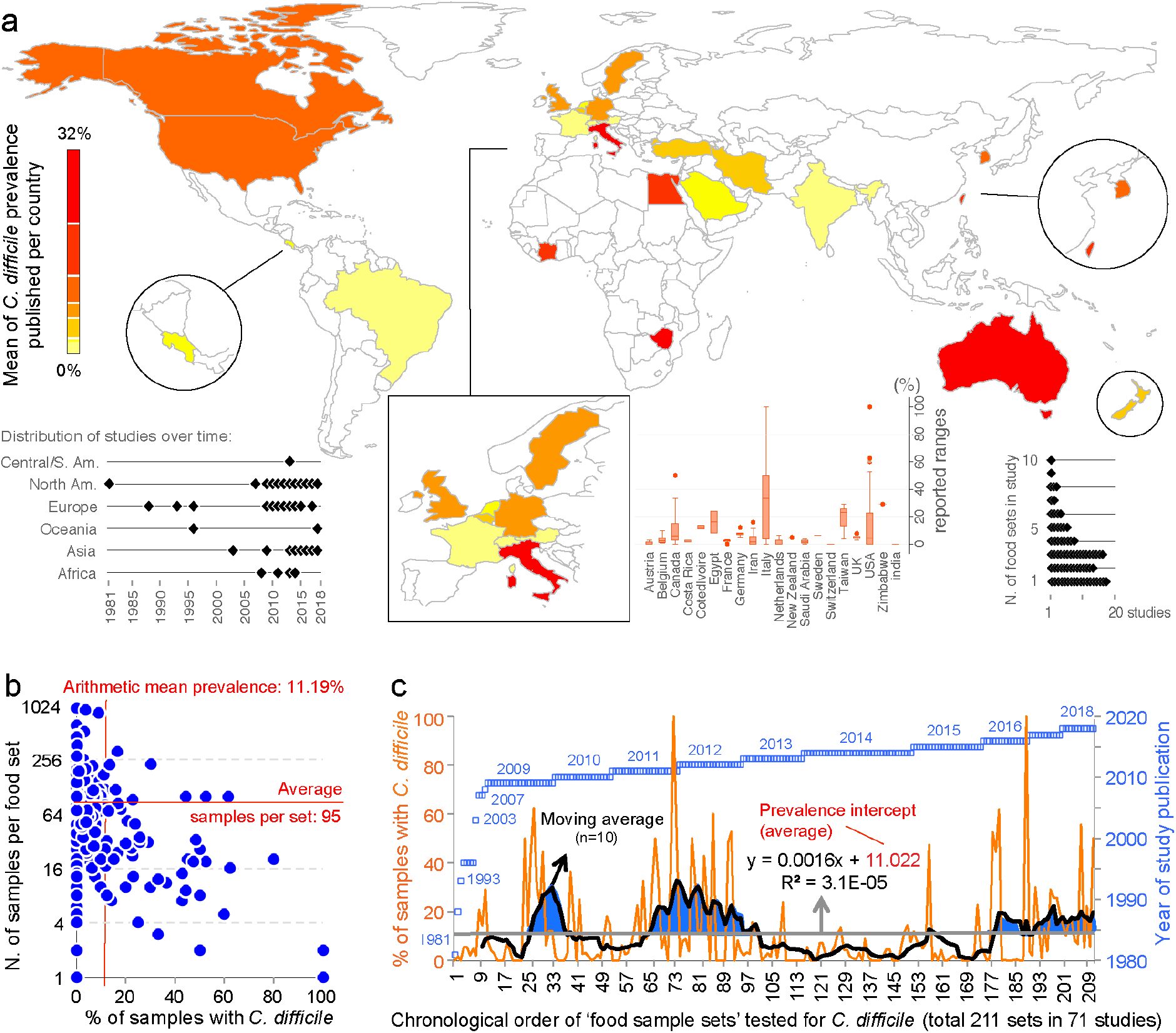
Global distribution and temporal oscillations of the reported prevalence of *C. difficile* in the human diet. Includes publications between 1981 and 2018. **a**) Distribution of studies included in the meta-analysis (n=79) with mean prevalence per country. **b**) Scatterplot (correlation) of number of food samples processed (sample size) for each food set sample and the percentage of samples with *C. difficile*. Insets, Example of prevalence variability among food items tested per country, distribution of studies over time, and number of food item categories (eg., beef, vegetables, poultry) tested per study. **b**) Chronological order of reported proportions for various food samples tested as sets irrespective of food category (food sample sets, n=230). Note that some studies collected samples from >1 food item category for culture of *C. difficile*.

Historically, the first study attempting to quantify the prevalence of CD in ready-to-eat foods was published by Fekety et al., in 1981 ^42^ in a hospital setting. Using a direct culture approach (effective for isolation of CD from environmental surfaces) on hospital meals, this study yielded no CD. The following year, two reports highlighted the potential foodborne and zoonotic potential of CD transmission to humans (Borriello *et al*.,1982 and 1983)^21,115^ but a period of quiescence lasted until 1996, when Broda *et al*.^116^ made a food science report of incidental isolation of CD from spoiled ‘blown-packed’ meats in New Zealand. Google citation statistics of Broda’s publication indicate that her findings were only relevant to food spoilage studies, and not cited on ‘public health’ or ‘food safety’ reports due to human health concerns until a report in 2006 discovered the presence of hypervirulent epidemic CD strains in food-producing animals and retail beef in Canada ^5,6^. No citations of Broda *et al*. occurred on the basis of foodborne/health concerns between 1996-2006 (0 vs. 30 citations on meat spoilage), but steadily increased to 27 foodborne citations after the 2006 reports ^5,6^ (51 citations on meat spoilage, mainly due to *Clostridium estercholaris*; Fisher’s p<0.001). Citation analyses support the reproducibility and historic context of our systematic review, with minimal publication interest on the ‘foodborne potential of *C. difficile’* before 2006. See **Figure 4** for a graphical representation of the historical context and order in which numerous laboratories around the world tested food items intended for human consumption since 1981.

**Figure 4.**
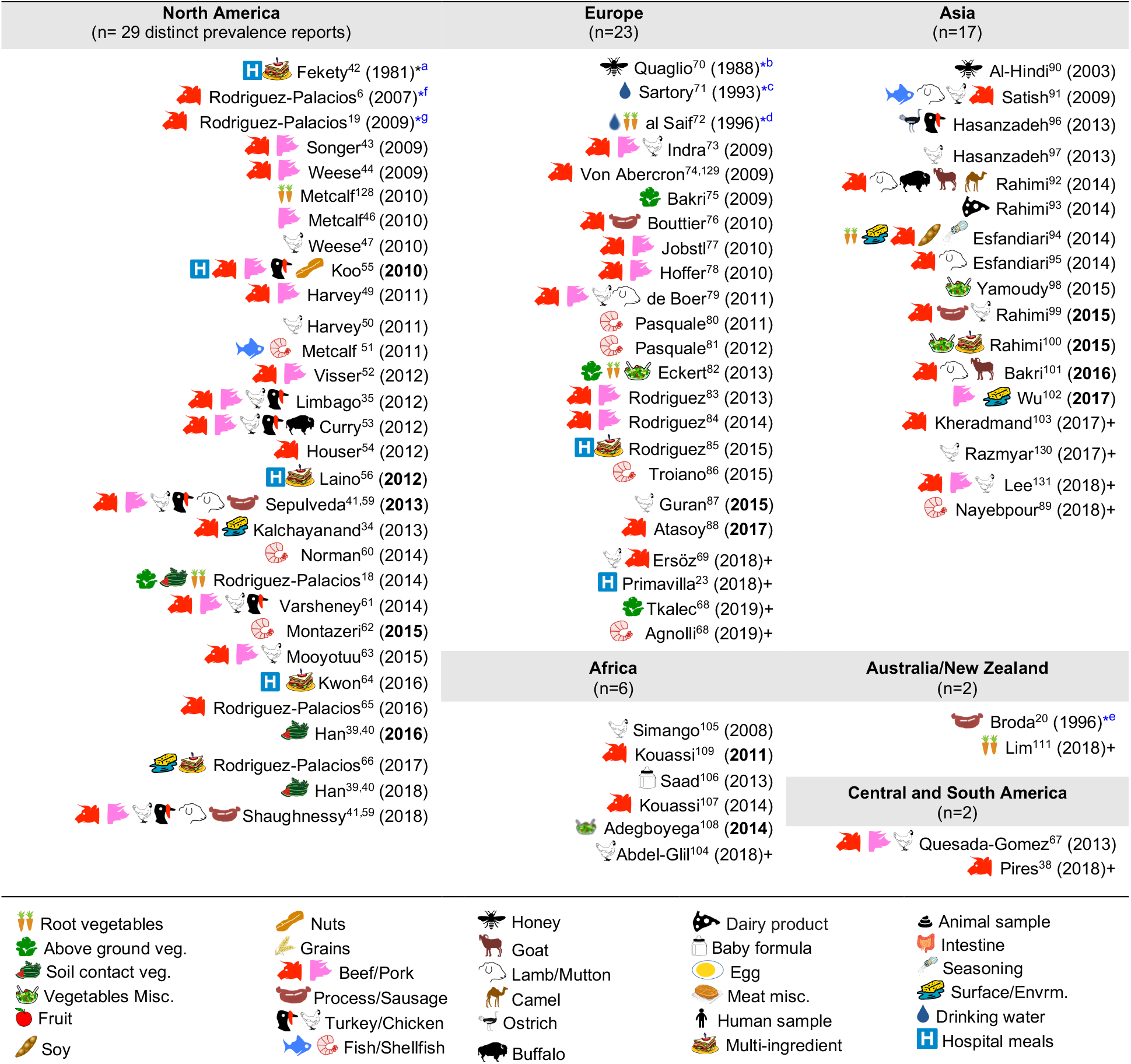
Graphical and chronological overview of food item categories (types) for human consumption tested for *C. difficile*: (1981-2019). Contextualization of food items tested in America, ^6,18,19,34,35,38-67^, Europe, ^23,68-88^ Asia, ^89-103^ Africa, ^104-109^ and Oceania,^20,110,111^. For a historic narrative see Results section. *First studies relevant to risk of ingestion *C. difficile* and food microbial safety epidemiology. ^a^First study in human hospital menus; negative results. Others ^55,56,64^ yielded positive results ^132^. ^b^First isolation from food produced by invertebrate insects – honey. ^c^First study in drinking water. ^d^First isolation of *C. difficile* from retail raw root vegetables. ^e^First isolation of *C. difficile* from animal-derived meat product, incidental finding while studying clostridia in spoiled and blown vacuumed packed sausages. No recognition of relevance to human health. ^f^First study on retail food derived from farm animals destined for mass scale production of food for humans with genotyping evidence of *C. difficile* hyper-virulent strains present in retail foods. Isolates obtained from retail ground beef purchased in Guelph, Ontario, Canada, 2004-2005. PCR ribotypes had assigned international nomenclature by Dr. Jon Brazier, U. of Wales, UK. ^g^First national systematic sampling study reporting seasonality of *C. difficile* in foods, Canada, 2006.

Although the cumulative prevalence of CD in the foods tested has been 4.1% globally at the study-level (two-tail 95%CI=-3.71, 11.91, **Table 1**), we demonstrate that the cumulative prevalence has distinct patterns of heterogeneity (variance) depending on the region, being comparably lower at the study-level in Europe (1.9%; 95%CI= −7.49, 11.29; see **Figure 5** for cumulative estimates in other regions).

**Figure 5.**
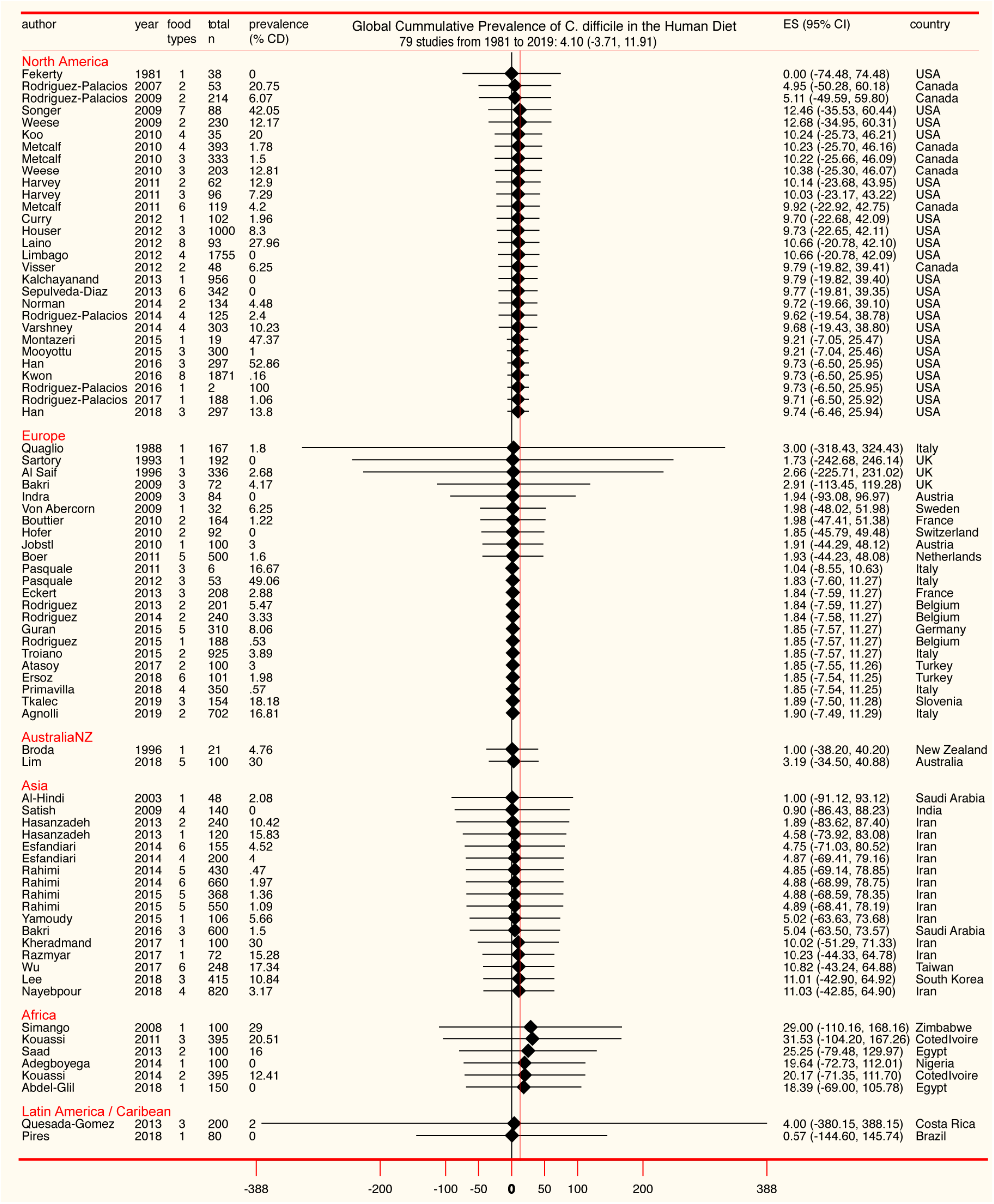
Forest plot of cumulative prevalence of C. difficile in foods for studies published since 1981, in chronological order per region (‘study-level’, n=79). Notice heterogeneity across regions. The detailed decomposed heterogeneity at the ‘food-item-category’ level (food types), for 232 food sets is presented below.

### Overall Food Contamination: Food-type level analysis

Because most studies (>75%) tested more than one ‘food-item type/category’ (*e.g*., ‘beef’, ‘vegetables’; 2.95±1.8 categories/study), and because pooling data from distinct food categories as a single CD prevalence for each study was deemed biologically inappropriate, and non-informative to generate food-based risk ranks, we extracted data separately for each food item tested in all studies. Thus, together, this meta-analysis represents 21,886 samples of retail foods tested across 230 ‘food item sample sets’. On average, each food set comprised 92±127 samples; maximum=956. For the pooled analysis, the 232 food sets were grouped into 20 food categories (*e.g*., ‘pork’, ‘seafood’, ‘mixed meats’), being ‘beef’ the most studied commodity (see cumulative statistics in **Supplementary Table 3)**. Reported CD prevalence at the ‘food-category level’ ranged from 0 to 100%.

As a single unweighted statistic, the arithmetic mean for the CD prevalence in foods at the food-type level was 10.6±16.6% (**Supplementary Table 4**). Because differences exist across regions and food tested categories, and because estimations depend on the inclusion of data from zero prevalence studies, we then computed the overall adjusted weighted meta-analysis cumulative prevalence considering the sample sets and regions, and three statistical methods to account for the 0% prevalence in CD-negative studies. Notice that **Figure 6** illustrates the heterogeneity (I^2^ statistics) across regions and the 230 food sets, at the same time it illustrates that the overall of *C difficile* in foods ranges between 4.5% (95%CI=3-6%, for all CD-positive and CD-negative studies combined) and 8% (95%CI=7-8%, for the CD-positive studies only).

**Figure 6.**
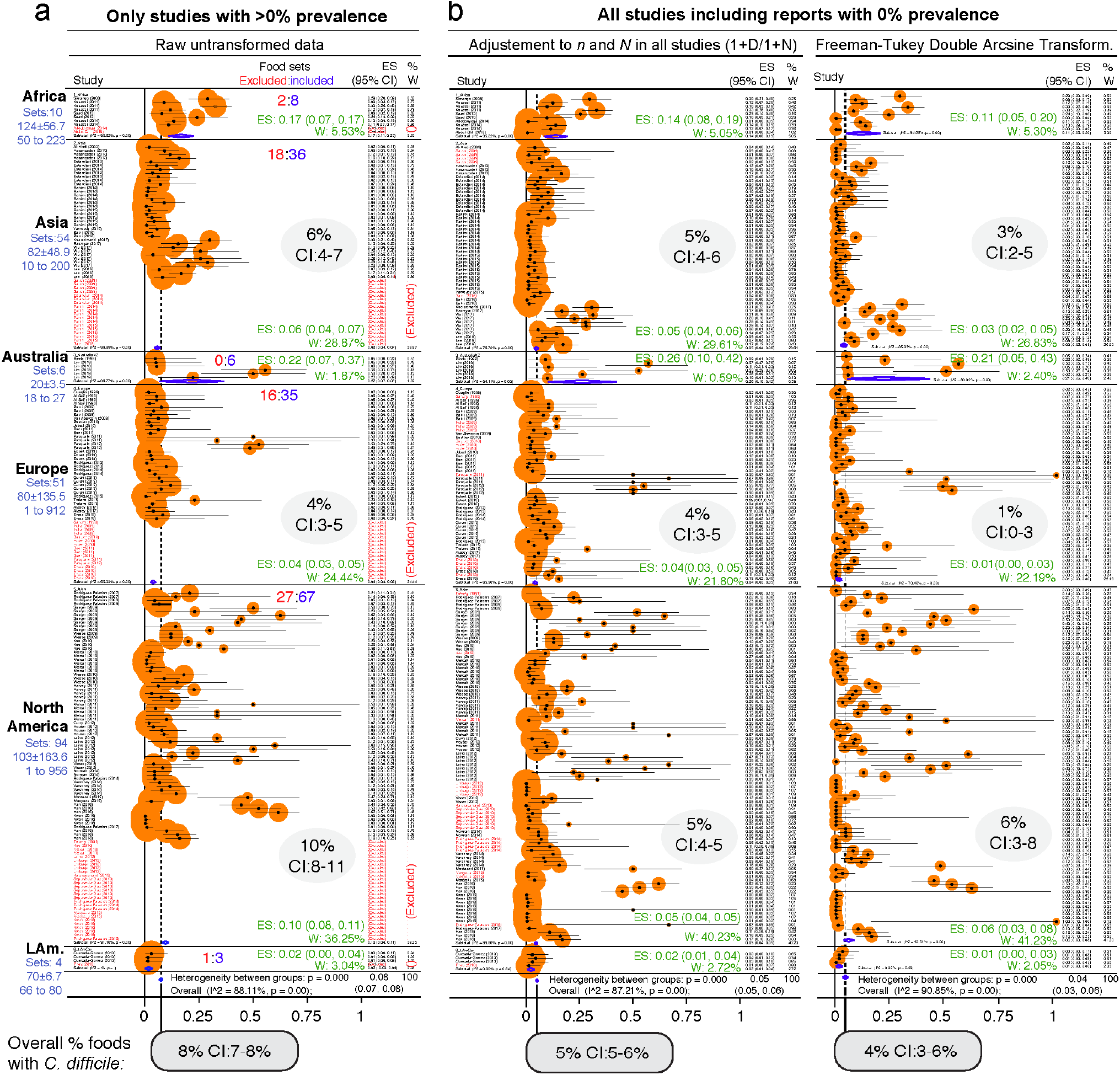
Forest plot of weighed prevalence of *C. difficile* at ‘Food-category’ level. (‘food item sets’, n=232). A version in PDF that can be magnified to high resolution is available in FigShare. Each plot represents different analytical strategies that differed on the method used for data transformation to deal with ‘zero’ prevalence reports. Data ranked by author and year. See estimates (ES) and weights (W) for each region in green and shaded ovals. Note that the confidence intervals (CI) overlap irrespective of analytical adjustments. **a**) Meta-analysis conducted with untransformed proportions. This mathematically excludes food sets with 0% prevalence (red font, approx. 25%). **b**) Meta-analysis conducted after adding 1 to the denominator and numerator (n/N), and after using the Arcsine Transformation of the raw data which forces the inclusion of adjusted data derived from ‘zero’ prevalence. Note the ranking of studies excluded in plot panel a (red font), are re-ranked in adjusted analyses. The smaller size of circles with the arcsine transformation illustrates better adjustment of heterogeneity. Confidence intervals are exact binomial (Clopper-Pearson). P<0.05 indicates pooled prevalence is different from zero. I^2^, heterogeneity test, p<0.05 indicates the ‘true effect’ across studies is not the same. Random-effects, DerSimonian/Laird statistics.

### Heterogeneity and Overall Prevalence of *C. difficile* in Foods is Independent of Culture Method

To date, one of the most cited factors to explain differences in CD across food studies is the existence of variability across methods and reagents (**Supplementary Table 5**). Although we have not seen recovery differences for antibiotics used as selective reagents in food studies (cycloserine-cefoxitin vs. cysteine hydrochloride-norfloxacin-moxalactam, CDMN)^**11719**^, we examined the role that culture methods play in this meta-analysis.

Although studies clearly report the use of anaerobic jars ^23^, culture media (e.g., CDMN, BHI), and homogenization methods for sample disruption (*e.g*., stomachers, blenders) which mix samples with room air, unfortunately, most studies did not specify clearly if reagents were pre-reduced (incubated anaerobically prior to utilization) or if protocols were anaerobic ^38,69,104^. Because 73.4% of studies did not use positive controls (58/79; **Supplementary Table 6**), it is impossible to infer if protocols were fully anaerobic. To test if the incubation of CD spores in non-reduced media (*e.g*., agar freshly removed from refrigerator) inhibits CD recovery, we conducted experiments *in vitro*. Using 1-year-aged spores from human PCR-ribotypes 078, 027, 077, strains 630 and ATCC 1869 ^7,36,65^, we observed that the use of non-reduced agars results in no CD recovery compared to using agars pre-reduced in an anaerobic chamber 4 hours prior inoculation (0/10 vs. 10/10, Fisher exact p<0.001). Because 26.9% of studies also reported short periods of incubation (*e.g*., overnight), we determined if short incubation influenced CD recovery. Of relevance, aged CD spores grew slowly requiring ∼72 hours to produce the same surface biomass (per colony on agar) as the produced by vegetative cells in 24 hours. Although results indicate that nonreduced media and short incubations could yield negative results, we deemed these to be common error factors randomly distributed across methods.

Thus, we next examined the role of overall culture strategies, by cataloguing and grouping all reported methods into six different categories based on sequence of isolation steps and three culture strategies: *i)* direct plating on agar, *ii)* enrichment of the foods using liquid media prior to culture on agar, and *ii)* the use of ethanol or heat to eliminate non-spore forming microbes in foods prior to culture in liquid media or agar to favor the growth of CD spores. Frequency analysis showed that almost three-quarters of all food samples tested (70.8%) used the methodological strategy reported in the first index report of CD in foods in 2006 ^**5,6**^. Confirming that the five remaining methods had comparable CD recovery, univariate and weighted predictive meta-analysis, showed that all the six methods were statistically similar (see **Figure 7**).

**Figure 7.**
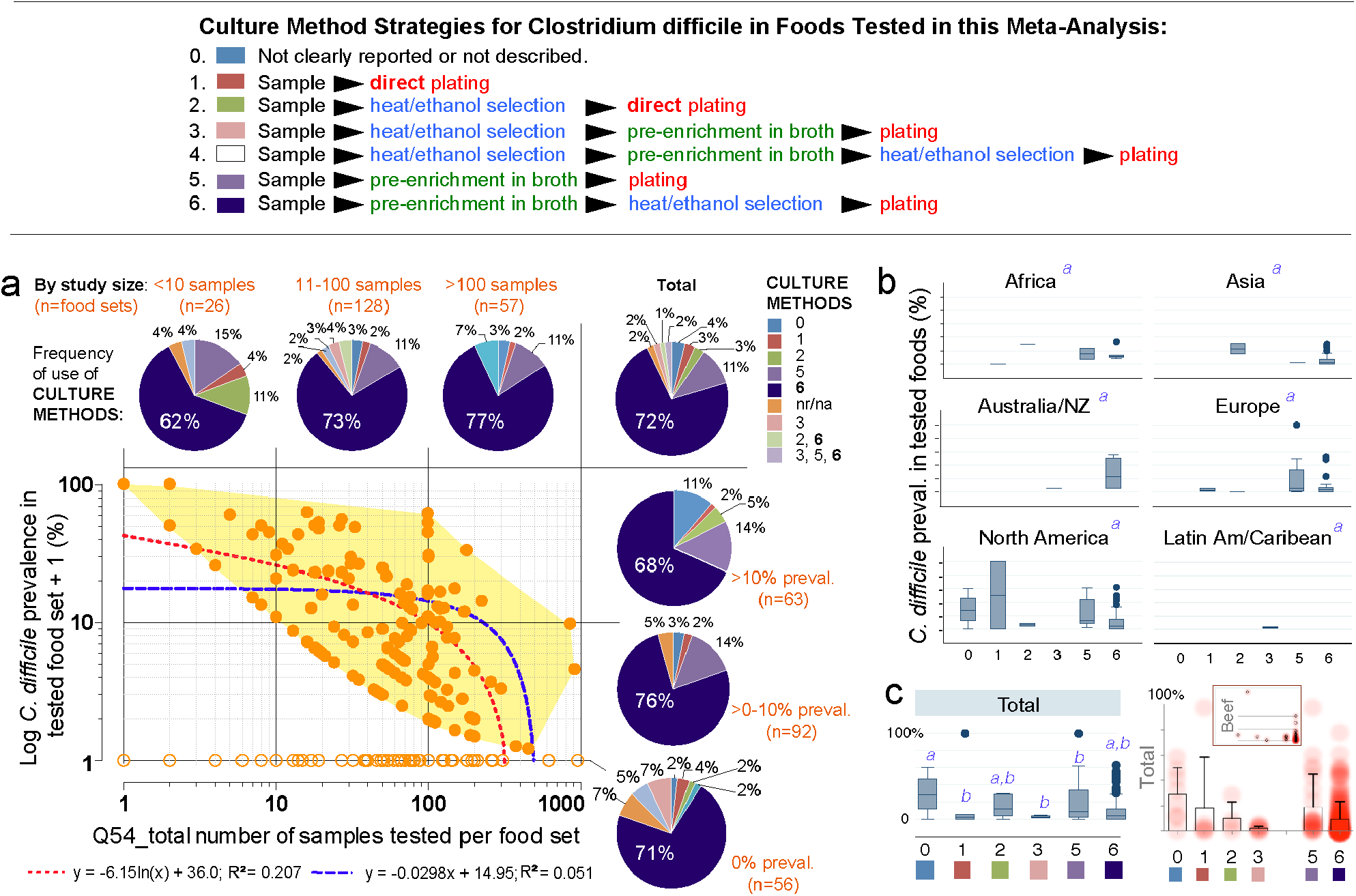
The isolation of *C. difficile* from foods is independent of the culture methods. **a**) Overview of culture sequence/strategies. **b**) Distribution of culture methods (pie charts) across the spectrum of study size or reported prevalences in this systematic review (scatter plot and best univariate linear fit models). Most studies used the enrichment method described by Rodriguez-Palacios *et al*., since fist studies reporting *C. difficile* in food-producing young animals, and retail beef ^5,6^ (dark blue in pie charts, #6). na/nr, not reported. **b**) Boxplots of reported prevalences across regions. Controlling for study name, geographical region, and culture method, there are no differences across regions in the log norm prevalence data in multivariable analysis (italic superscript ‘a’, adjusted p>0.1; generalized linear model: outcome, a99a_percentplus1log; categorical variables, a46a_overall_cult_apprch a9_pub_region a5_stydy_id). **c**) Cumulative standard boxplot and density scatter boxplot of reported prevalences across culture method. Inset, Density scatter boxplot for beef samples illustrates reproducibility of cumulative data. See statistical details in **Supplementary Table 5**.

Publication bias, journal impact factor, and the amount of food tested were also ruled out as sources of variability. However, we discovered that the number of samples tested per food set correlated inversely with CD prevalence (linear regression p=0.007; meta-regression p=0.067 controlling for region/method, **Supplementary Figures 1-2** and **Supplementary Table 7**). Although seasonality has yielded heterogeneity in food animals (*i.e*., low prevalence in summer; high in winter in temperate regions), seasonal variability could not be tested since 85.9% of studies did not include referents or surrogates for season. Together, **Figure 7** and the analysis described illustrates that different culture strategies cannot explain the prevalence heterogeneity reported in the literature, and confirmed that all studies can be integrated in this meta-analysis.

### Contamination Risk Analysis Ranks Vegetables and Seafoods as High-Risk Food items

Of relevance to risk statistics, over one-quarter of food sets were CD negative (64/232; 27.8%, 95%CI= 22.2, 32.2). However, from a clinical perspective, doctors and patients could benefit by knowing which foods are more likely to be contaminated to determine how diets can be adjusted during periods of increased susceptibility (*e.g*., cancer, IBD). For instance, by cooking or avoiding high-risk foods.

Since different food items could be contaminated with different probability risks, we calculated risk ratios (RR) to rank each food group with respect to the food yielding the lowest combined prevalence of CD, and also using meta-analysis weighted estimates (**Figure 8**). Using milk as a reference (which had the lowest prevalence, but clinically important CD strains) ^22^, vegetables, seafoods and pork had the highest RR. Compared to milk, vegetables were 21.9 times more likely to yield CD, while seafoods and pork were 14.3 and 12.9 times more likely, respectively. **Figures 9, 10, 11**, and **12** comparatively illustrate the weighted prevalences for the following food categories: beef and vegetables, poultry, pork and seafood. Comparing retail beef, leafy-green vegetables and root vegetables, **Figure 9** illustrates that leafy green vegetables are twice more as likely to carry CD compared to root vegetables. **Supplementary Figures 3-5** display detailed forest plots for both statistical methods for beef and vegetables, and for all the food tests tested including mixed meals, and others based on the biological origin (animal/plant) of the food.

**Figure 8.**
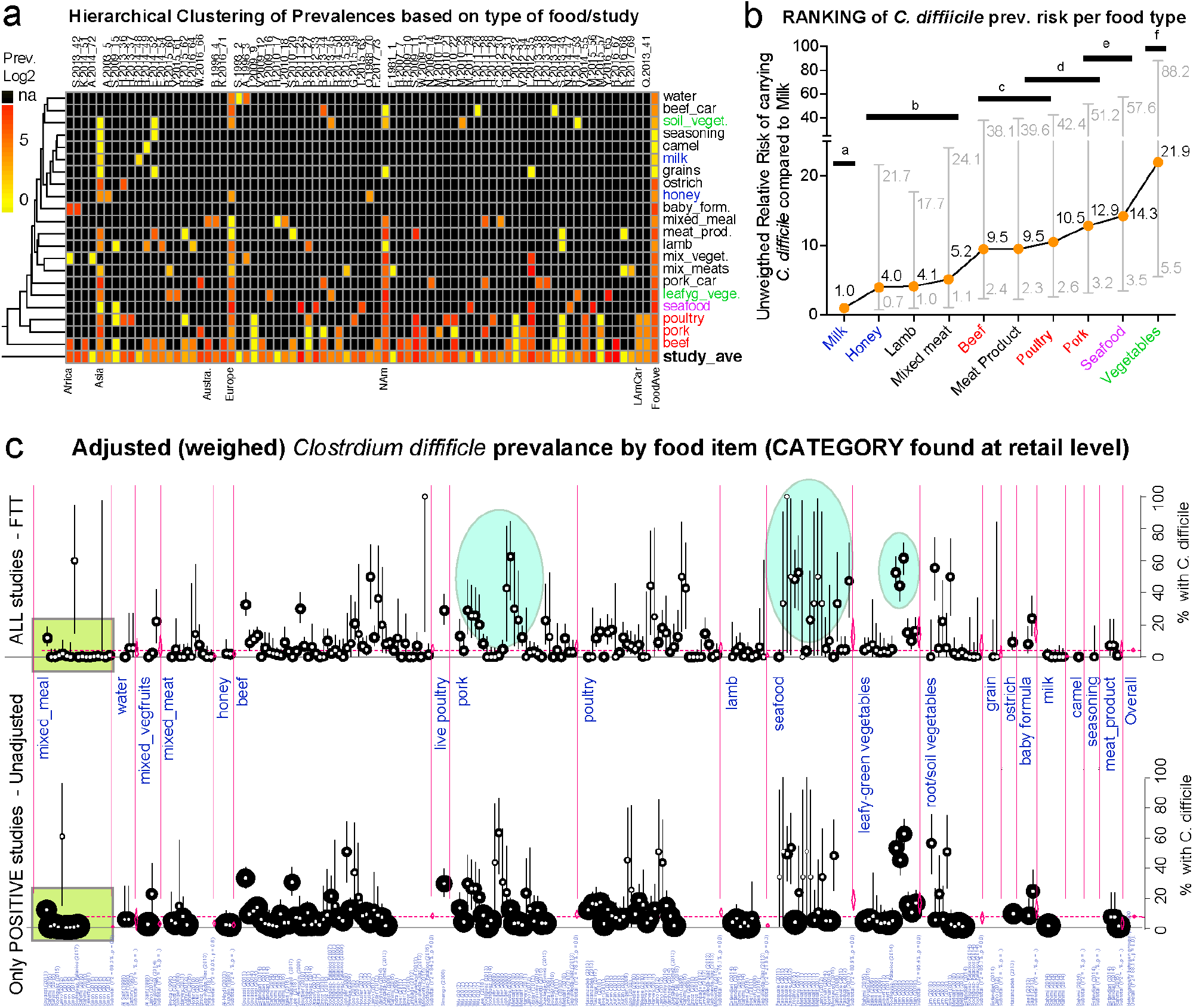
Global risk ranking of foods based on probability of carrying *C. difficile* illustrates higher heterogeneity for pork, poultry, seafood and vegetables. A version in PDF that can be magnified to limiteless resolution is available in FigShare. **a**) Hierarchical unsupervised analysis of reported prevalence of *C. difficile* (CD) in various food items (y-axis labels) across regions and studies (x-axis). Note that several studies processed various types of foods. na, not tested. **b**) Ranking of foods based on the expected risk of carrying CD (Relative Risk Ratios [RRR] and 95% CI). Note that the RRR ranks beef, poultry, pork, and vegetables at different levels although they cluster together in panel a, which clusters these products together because those were more commonly tested across regions. Horizontal bars connect products with statistically similar RRR. Distinct superscripts denote statistical differences, Chi-square p<0.05. **c**) Meta-analytic display of weighed prevalence estimates at the ‘food set’ level. Top panel displays data from all studies, including ‘zero’ prevalence reports (Arcsine transformation, homogeneous adjustment for variability, see comparably-sized small circles), while the top panel displays data of only positive studies (larger variably-sized circles, see area within rectangular polygon). Vertical ovals in top panel highlight representative clusters of reports describing high prevalence of *C. difficile* in certain foods, supports raking statistics in Panel b. Leafy green vegetables are ranked high since estimates are from studies with larger sample sizes/more weighed influence (small circles, narrower CI in bottom panel; larger circles in arcsine-adjusted top panel).

**Figure 9.**
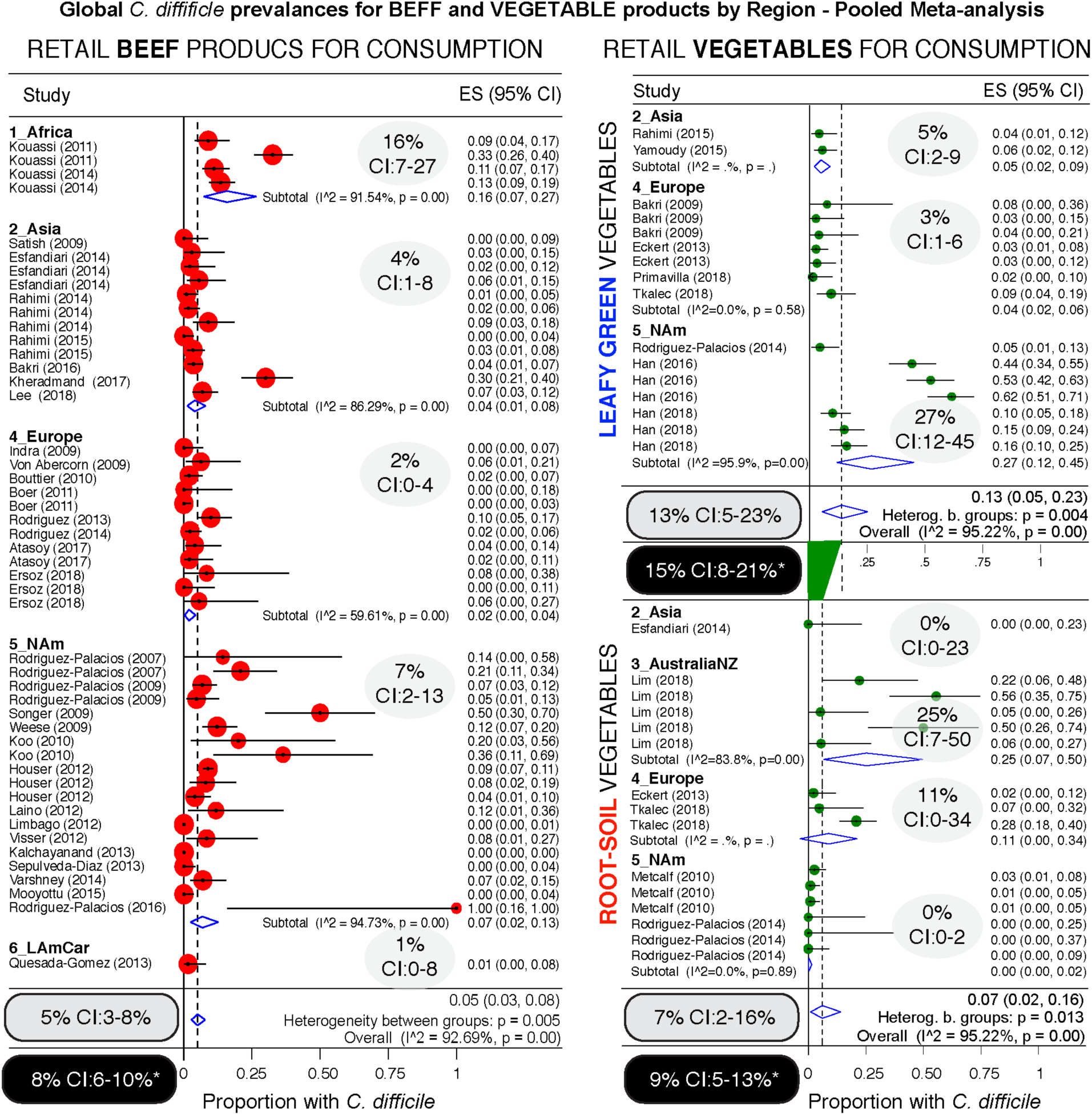
Comparative global and regional prevalence of *C. difficile* in beef and vegetables. Forest plots using the Arcsine Transformation of the raw data force the inclusion of adjusted data derived from ‘zero’ prevalence studies. Confidence intervals (CI) are exact binomial. Rectangular ovals denote overall estimates. Shaded ovals, region estimates. Back ovals denote overall estimates from unadjusted meta-analysis (detailed plots with higher prevalence estimates from unadjusted data to include only CD positive studies are in **Supplementary Figures 3** and **4**). Note larger variability among studies conducted with vegetables (wide overall CIs) when compared to variability for beef products (narrow overall CI). Leafy green vegetables are twice more commonly found to contain CD compared to root vegetables. Analysis of these three food type categories, based on weighed mean prevalences, ranks leafy green vegetables as more likely to carry CD, and beef products the least likely.

**Figure 10.**
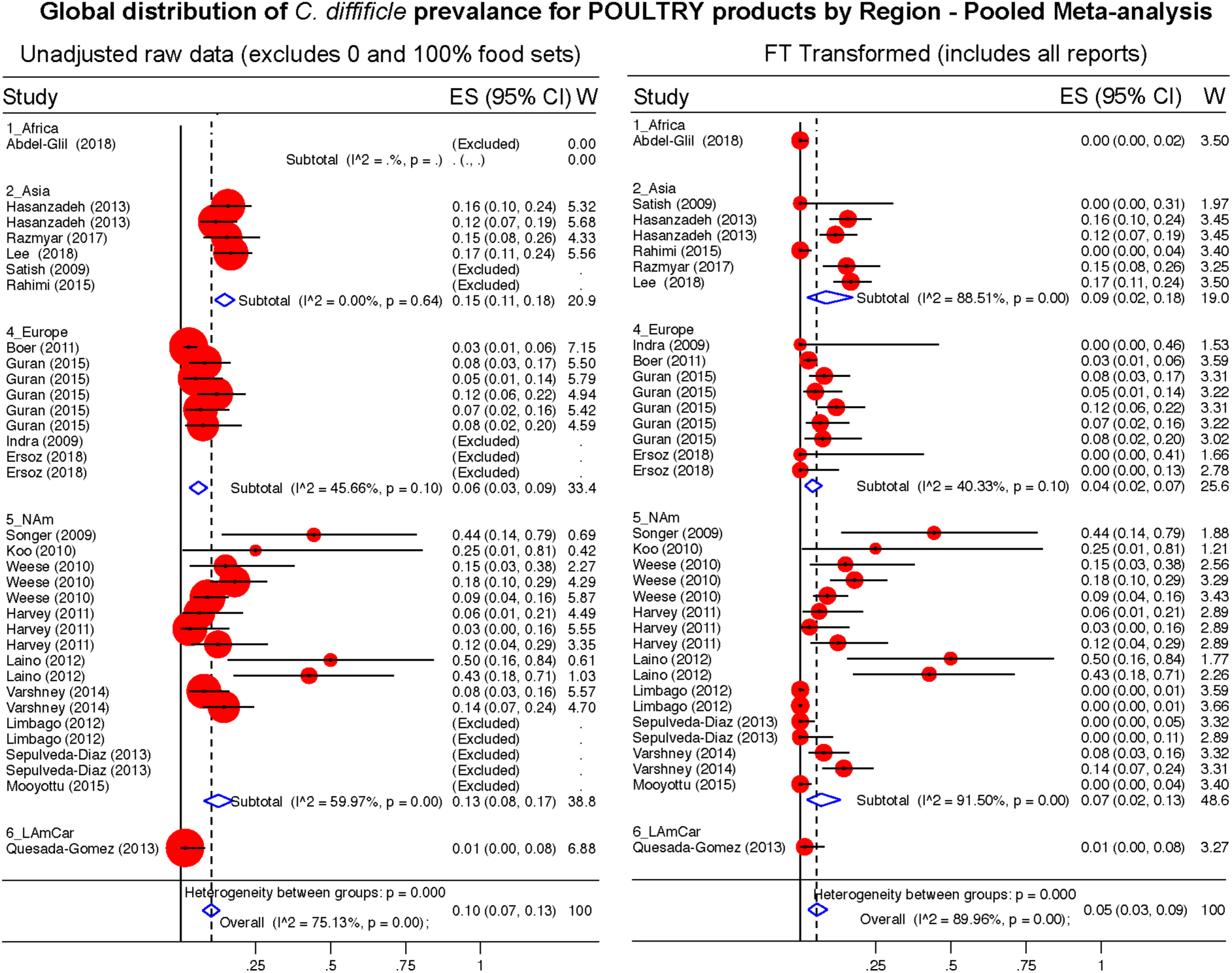
Global Prevalence of *C. difficile* in Poultry. Untransformed data (left panel) and Freeman-Tukey Double Arcsine Transformation (right panel).

**Figure 11.**
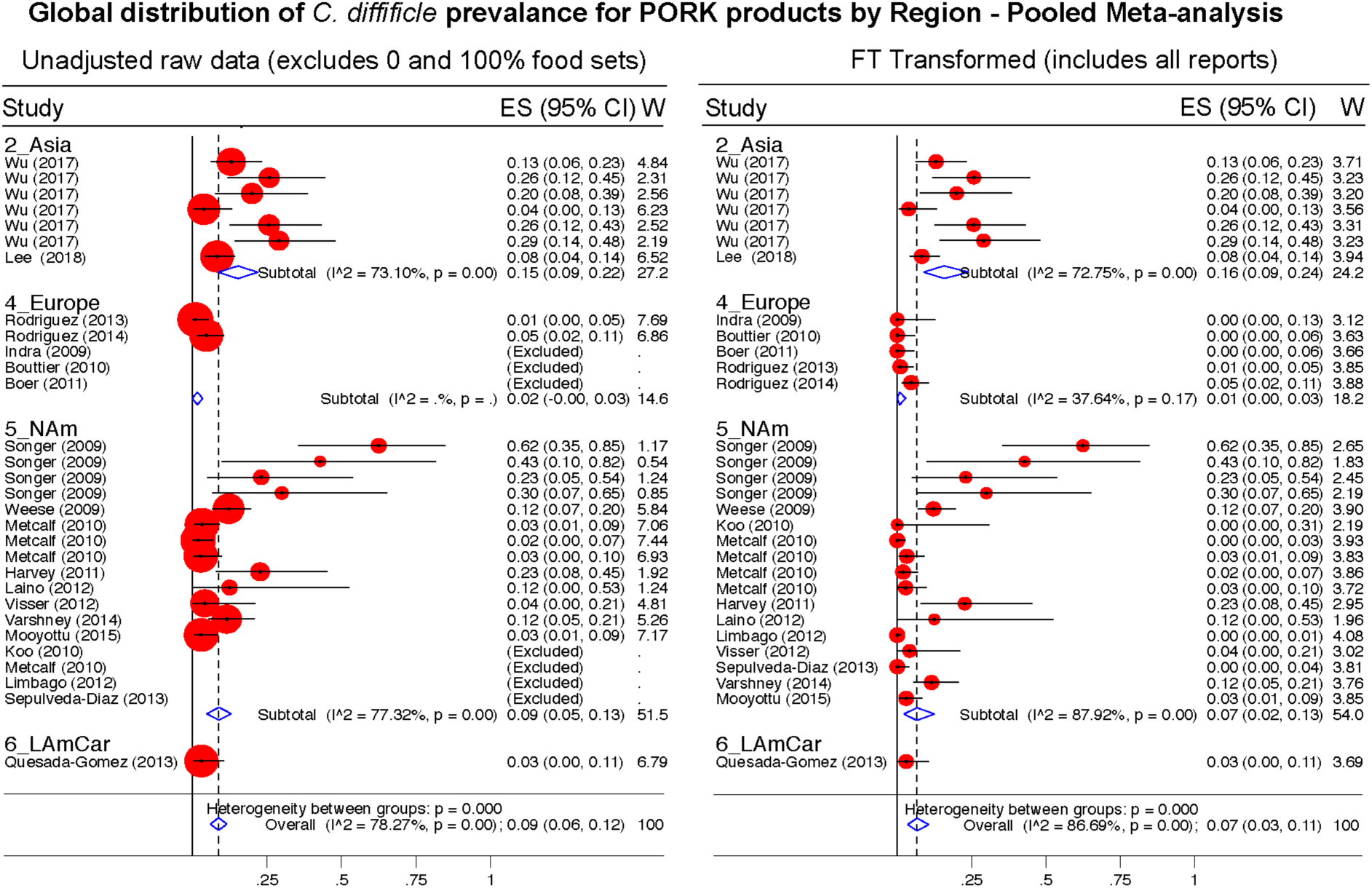
Global Prevalence of *C. difficile* in Pork. Untransformed data (left panel) and Freeman-Tukey Double Arcsine Transformation (right panel).

**Figure 12.**
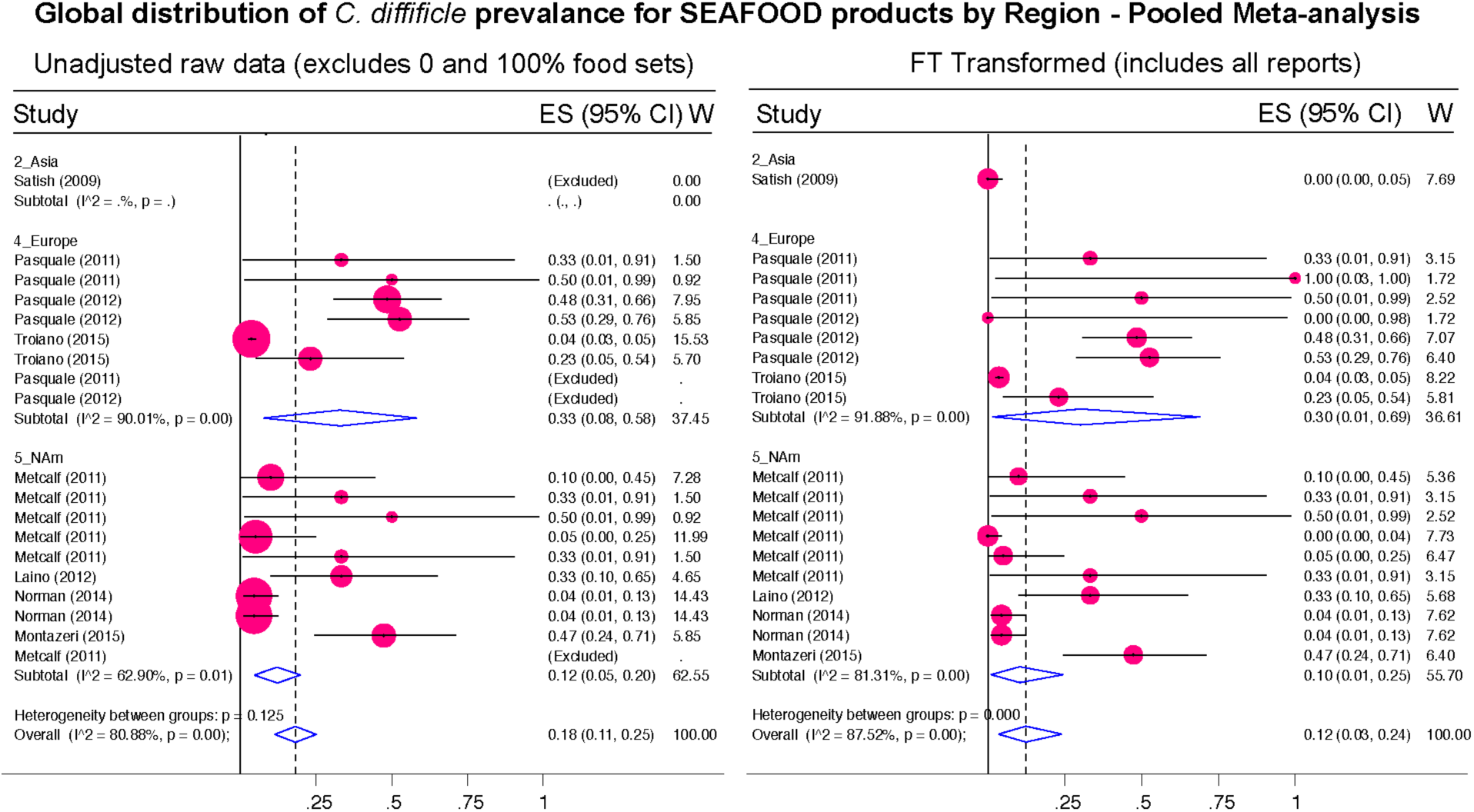
Global Prevalence of *C. difficile* in Seafood. Untransformed data (left panel) and Freeman-Tukey Double Arcsine Transformation (right panel).

### The Probability of Recovery *C. difficile* from Foods Increased Latitudinally Towards the Tropic

To determine whether the prevalence of CD in foods was influenced by Earth’s latitude, we added the positional coordinates to the dataset. Both unadjusted and arcsine adjusted meta-regression revealed that latitude determines the magnitude by which CD has been isolated from foods worldwide. While longitude was nonsignificant, latitude had a negative linear correlation with CD prevalence (in a *y* = β_0_ + β_1_χ_1_ model; **Figure 13a**). Since several studies reporting high prevalence were from mid-range latitudes, collectively the data displayed a concave pattern (in a *y* = β_0_ + β_1_χ_1_ + β_2_χ_1_^2^ model; **Supplementary Figures 6-7**). However, after dividing the 230 food sets into 22 food-per-continent subsets to control for longitude (*e.g*., beef in Africa vs. Asia), regression slope analysis (in *y* = β_0_ + β_1_χ_1_) showed that latitude negatively correlation with prevalence in 94.5% of the 22 data subsets (Sign p<0.0001). Such reproducible correlation was not due to chance, since random allocation of latitude values in 25 simulations showed nonreproducible slopes (Sign p=0.35). Findings were also validated using predictive spatial density map simulations on a 2D-plot representing the Earth’s surface (**Figure 13b-e**, adjusted p<0.001). Contour-density plots illustrate that the patterns of CD have a spatial latitudinal structure that is different from simulations of randomly spaced studies. For the first time, the prevalence of CD in the human diet is shown to have a latitudinal pattern over the Earth’s latitude. Occurring reproducibly across continental longitudes, with comparatively higher CD prevalence in regions closer to the tropic, this CD-in-foods trend is opposite to what is expected for CDI in humans, where most cases seem to occur more often (in temperate regions) away from the tropic.

**Figure 13.**
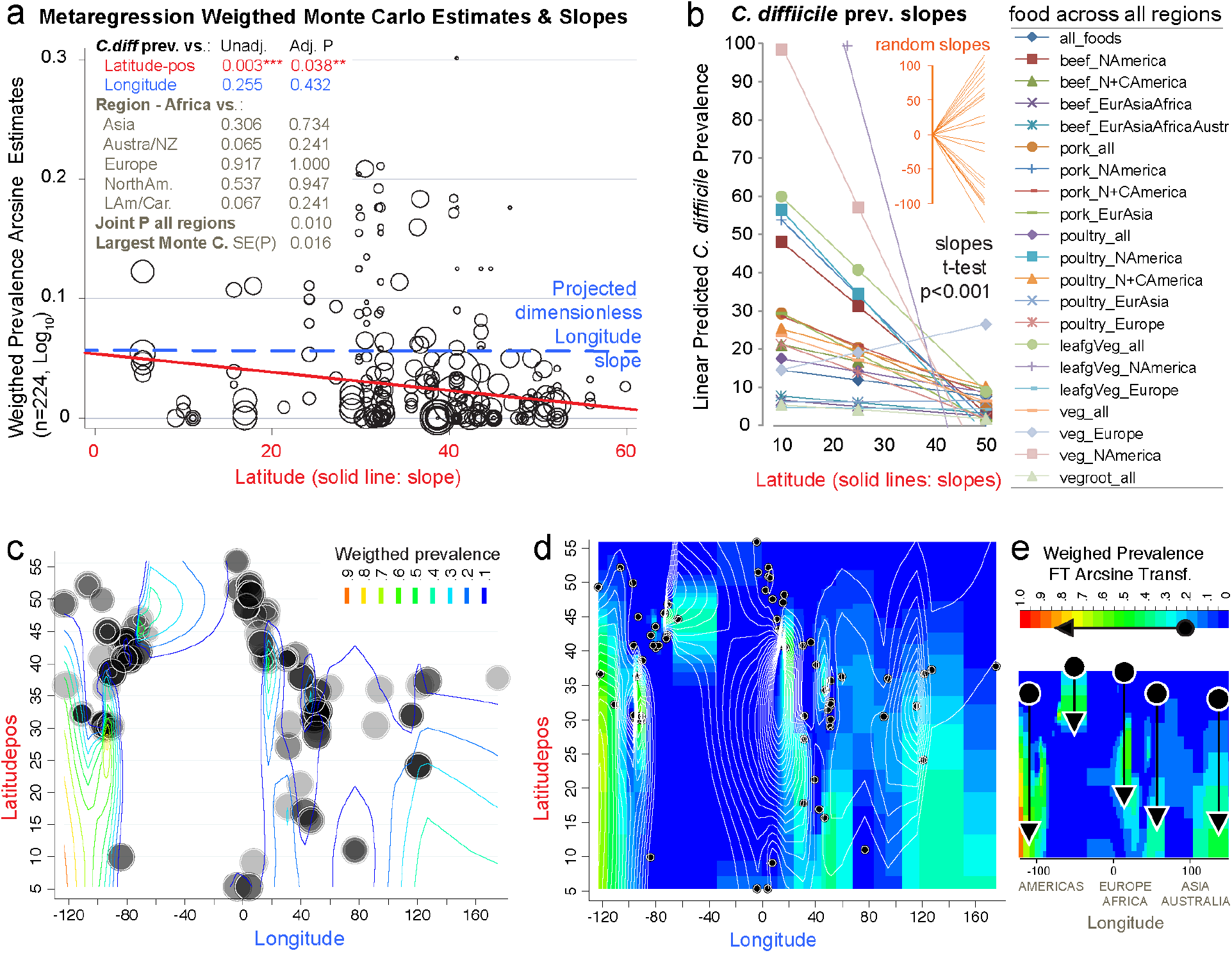
The probability of recovering *C. difficile* from foods increases towards the tropic. Linear correlation estimates, various meta-regression analyses controlling for confounders, contour plot simulation and Monte Carlo permutation test (n=224, 1000 perm, joint P=0.01) statistics revealed that latitude has been one of the most influential variables determining the magnitude and frequency by which C. difficile has been found in the human diet. **a**) Moment-based estimate of between-food-set study variance and display of weighed correlation between prevalence and the absolute latitude. Without Knapp & Hartung modification to standard errors. P-values unadjusted and adjusted for multiple testing. Note that longitude is not significant variable. **b**) Plot of linear trends derived from fitting linear models to actual data segregated by type of food and continents/regions aligned over distinct longitude ranges. Notice that except one slope, published reports have documented an inversed latitudinal trend. **c**) Contour line plot simulation of the weighed CD prevalence for all food items over absolute latitude and real longitude plane (semi-transparent circles of different sizes, the larger the circle, the greater the influence on overall simulation). **d**) Contour density and line plot simulation to help visualize the low prevalence estimates (near zero=blue) and latitudinal trends. Circles represent the location of the research centers were the studies were conducted or the centroid for the region that was sampled. **e**) Contour density simulation to illustrate that latitudinal trends (arrows) can cover different latitudinal ranges, depending on the region (e.g., short high arrow corresponds to Europe). In iterative simulations, it is to note that such density latitudinal trends tend to cluster between two extreme arrangement patterns but that the significance is independent of the region (**Supplementary Figures 6-7** for further details and statistics).

## Discussion

The present meta-analysis, for the first time summarizes the distribution of CD in the human diet, which we derived from data from 79 studies conducted between 1981 and 2019. Estimated various regional and global prevalence of CD in foods ranged between 3 and 8% globally, or between 0 and 22% regionally. We also identified for the first time a latitudinal trend in foods with increased rates of CD recovery in food towards the tropic. The analysis of almost twenty-two thousand samples across the globe, as a robust representation of the human diet, indicates that prevalence heterogeneity exists independently of culture methods. While study variability has been assumed to be consequence of culture method differences, our analysis (verified using I^2^ statistics) demonstrates that there were no significant differences for the CD prevalence across methods, and that most studies used the same methodology. Prevalence estimates also varied within studies conducted by the same author, which cannot be explained by variations in culture methods. Often, the same method was applied to different food items yielded different rates of CD contamination under the same report. Such differences reflect real variance of CD in the food supply.

Although earlier articles speculated that the identification of CD in foods could have been due to poor techniques and cross-contamination, high-quality studies have shown that contamination is an obsolete argument to discount the value of identifying toxigenic and even emerging virulent strains of CD in the food supply, which have been shown to be genetically similar to strains of clinical relevance in distant regions^7^.

Because a number of studies reported 0% of CD, it is possible that there are natural sources of contamination heterogeneity in foods, similarly to other known foodborne pathogens. There is substantive evidence to support that the risk changes as a function of climate, and latitude. It has been established that the tropic has ecologically greater microbial diversity ^127^, but how such diversity could determine the presence of *C. difficile* in the food supply across regions is uncertain. If CD contamination is higher toward lower latitudes, possible explanations could include that more diverse microbiomes in the gut, environment, ^127^ and foods towards the tropic could prevent CD colonization and CDI, since CDIs are more often reported in temperate latitudes.

Our study only examined the reported prevalence of CD in food items, regardless of the toxinogenic potential of the identified isolates, assessed on culture cells or in susceptible hosts. Virtually, every study recovering CD have determined that the isolates have had at least one of the three toxins or genes needed to fulfill the criteria for CD toxigenicity (*tcdA, tcdB, cdtA/B*). Similarly, numerous studies have used molecular methods to determine the epidemiological distribution of the isolates in human hospitals. However, because the performance, acceptability, and generalizability of molecular typing methods vary across regions, and because there is no a single unified system for CD strain typing or nomenclature worldwide to make meaningful comparisons at global scales, we refer the readers to the original publications to examine the strength of the genomic evidence reported in each epidemiological study. As historically highlighted, we emphasize that there is molecular evidence that the presence of CD in the human diet is genuine and not due to laboratory cross contamination with CD from human specimens. Major examples include the complete genome sequence of the first food derived PCR-ribotype 078 isolates from foods in Canada that matched contemporary strains affecting humans in the UK, in the mid 2000s, when there was no physical connection between the laboratories that reported both studies^7^. Supporting the remarkable risk for CD exposure via seafoods, we also highlight the latest report of CD in foods conducted in the Adriatic Sea where mussels and clams contaminated at a mean prevalence of 16.9% (CI: 14.1%–19.8%) carried a large proportion of CD representing diverse genotypes commonly isolated in European hospitals (113 CD isolates represented 53 genotypes, with 40.7% of them belonging to CD seen in CDI in hospitals).^133^

Although the present meta-analysis showed significant latitudinal heterogeneity, one of the limitations of the reported studies, and therefore the coordinate data used for the analysis, is that the latitudinal positioning of the samples collected and processed by each of the study authors is inferred for each research center, and it is not the actual coordinates of origin of each sample which was not reported in any study. However, since the analysis is conducted at the global scale and it is considered to be a proxy for the exposure risk, for all studies, which is relevant for the local communities in the districts sampled by the researchers, the analysis and findings are deemed pertinent and good indicators of the effect of latitudinal positioning and the CD prevalence trend observed at the global scale. Lastly, most studies have been conducted in Northern regions, however, to increase the study power we normalized all latitudes by squaring the latitudinal coordinates as the distance from the equator toward both hemispheres which is mathematically and geo-positionally standard method to gain statistical symmetry around latitude 0 (equator).

In summary, this study does not intend to make inferences/comparisons between north and south hemispheres. It only addresses the effect of absolute coordinates, which by the virtue of being positive (by squaring negative coordinates), they may be more representative or inflate the ecology in the north latitudes. Because local climates vary in opposing terms as latitude increases toward the poles (winter in north, summer in south, and vice versa, not controlled in this study because precise temporal referents were not reported in the reviewed studies) it is advisable that future studies provide databases containing the coordinates, day/month of the year and air temperature for each sample, and the CD test results to validate and further test the latitudinal trend and hypothesis herein generated in this systematic review.

In conclusion, it is reasonable to infer from our analysis that there is no a single number that summarizes the complexity of CD in the human diet worldwide. Until the dynamics of CD over space and time are better defined, doctors could advice patients and communities at risk to cook their meals better and give other simple suggestions, such as avoiding high-risk foods that are commonly consumed raw (*e.g*., fresh produce), until the patient’s susceptibility to CDI decreases. From a clinical and prevention perspective, patients could benefit by knowing which foods are more likely to be contaminated with CD to determine how to adjust their diets during periods of increased susceptibility. Considering that ∼10% of the samples in this study (∼20 grams per sample, over 1 overfilled tablespoon) were contaminated with CD, which represents only a fraction of an average meal size per person, it is possible that consumers are exposed to CD very frequently. If a person consumes 500 g of food per day, estimates could suggest than in average one table spoon full of meal in every 13 (260 grams of meal) could be contaminated with CD, if not cooked properly. Basic recommendations emphasizing food safety practices updated to CD (using >85°C for 10 minutes, or even better, boiling temperatures) ^65,66,118,119,120^, could prevent inadvertent exposure especially if patients are affected with debilitating conditions that increase the risk for CD intestinal colonization and infection. Future publications should include in their design and reporting descriptors for climate, ambient temperature, season and latitude.

## Data Availability

Data Availability and Open Access. All data, easily extractable from the manuscripts listed in the Supplementary References, will be made available by the corresponding authors upon reasonable request. This article is intended for publication as open access under the terms of a Creative Commons Attribution- International License.

## Acknowledgments

Study was conducted with internal funds allocated to the corresponding authors. SI is an Assistant professor of Human Nutrition and Food Microbiology, the official Food Safety State Specialist for the state of Ohio in the USA, and is a current Expert consultant on food safety for the Food Agricultural Organization of the United Nations. NB received support from the Fulbright as a senior scholar and visiting professor in mathematics at the University of North Carolina. ARP receives support from the National Institutes of Health via grants R21 and R01 subaward mechanism, and is the Director of the Germ Free and Gut Microbiome Core at the Digestive Diseases Research Institute and is the Technical Director of the Mouse Models Core at the NIH Silvio O Conte Cleveland Digestive Diseases Research Core Center.

